# Gender Affirming Hormone Therapy induces specific DNA methylation changes in blood

**DOI:** 10.1101/2021.07.05.21260016

**Authors:** Rebecca Shepherd, Ingrid Bretherton, Ken Pang, Anna Czajko, Bowon Kim, Amanda Vlahos, Jeffrey D. Zajac, Richard Saffery, Ada Cheung, Boris Novakovic

**Author notes:** Co-senior authors (Ada Cheung, Boris Novakovic).

## Abstract

DNA methylation is an epigenetic mark that is influenced by underlying genetic profile, environment, and aging. It also plays a key role in female-specific X-chromosome silencing in mammals. In addition to X-linked DNA methylation, sex-specific methylation patterns are widespread across autosomal chromosomes and can be present from birth or arise over time. In individuals where gender identity and sex assigned at birth are markedly and consistently incongruent, as in the case of transgender people, feminization or masculinization may be sought through gender affirming hormone therapy (GAHT). In this study we profiled genome-wide DNA methylation in blood of transgender women and transgender men, before and during GAHT (6 months and 12 months into hormone treatment). We identified several thousand differentially methylated CpG sites (DMPs) in both people undergoing feminizing and masculinizing hormone therapy, the vast majority of which were progressive changes over time. Sex-specific DNA methylation patterns established in early development are largely refractory to change in association with GAHT. The small number of sex-DMPs that were affected by GAHT were those that become sex-specific during the lifetime, known as sex-and-age DMPs.

## Introduction

In the era of personalised medicine, inclusivity of all genders and sexes in research is vital for providing equitable and inclusive healthcare (Moyer et al., 2019). Although often used interchangeably, gender and sex are separate constructs. ‘Gender’ refers to one’s self-perception of sex (often termed ‘gender identity’) while ‘sex’ refers to the underpinning biological features, which is assigned at birth and defined by reproductive anatomy or sex chromosomes (Dotto, 2019; Short et al., 2013). Gender dysphoria occurs when an individual’s gender identity and sex assigned at birth do not align. In those where these are markedly and consistently incongruent, as in the case of transgender individuals, feminization or masculinization may be sought through gender affirming hormone therapy (GAHT). In those seeking feminization, GAHT includes estrogen (in the presence of absence of progesterone) in combination with anti-androgens, while testosterone therapy is used for those seeking masculinization.

A wealth of research exists on ‘sex-specific’ immunity, including evidence of sex-specific vaccine and infection responses (reviewed in (Klein and Flanagan, 2016; Markle and Fish, 2014)). Additionally, sexual dimorphism exists within several immunological diseases, such as the heightened prevalence of some autoimmune and inflammatory diseases in among adult individuals presumed female at birth compared to adult individuals presumed male at birth (Selmi and Gershwin, 2019). However, these findings are based on binary comparisons based on sex, and it is important to broaden the current understanding of immunity to include transgender individuals. Moreover, sex hormones have an immunomodulatory role (reviewed in (Bereshchenko et al., 2018; Bouman et al., 2005; Shepherd et al., 2021)), and therefore the effects of GAHT warrant investigation as it marks a period of profound change in the internal hormonal milieu.

DNA methylation is an epigenetic mark that influences gene expression in a context-specific manner (Jones, 2012). This mark is highly dynamic during pre-implantation development and during cell differentiation (Smith et al., 2014), but also susceptible to environmental factors throughout the lifetime (Feinberg, 2018). The inactivation of the second X chromosome in females is an example of a co-ordinated sex-specific epigenetic remodelling process. This process involves a non-coding RNA, chromatin remodelling and finally the addition of DNA methylation on the inactive X (Heard et al., 1997). In addition to X-inactivation associated DNA methylation differences (Heard et al., 1997; Weber et al., 2007), sex-specific (XX females versus XY males) DNA methylation patterns on autosomal chromosomes have been reported in a range of cell types (Gabory et al., 2009; Hall et al., 2014; Inoshita et al., 2015a; Mamrut et al., 2015). At least some of these are potentially due to hormonal change, but these are hard to delineate from genetic influence, as hormones are regulated by biological sex in isolation. Periods of hormonal change have been shown to induce changes in DNA methylation over time, including pregnancy (Gruzieva et al., 2019b), puberty (Almstrup et al., 2016; Thompson et al., 2018), menopause and menopausal hormone therapy (Cheishvili et al., 2018; Ronkainen et al., 2010). Analysing GAHT-induced DNA methylation change provides a unique model to study the effect of hormones separate from genetics. Here we report the first Epigenome-wide association study (EWAS) of GAHT, and describe the longitudinal changes observed in the blood methylome of 13 transgender men and 13 transgender women newly commencing GAHT at baseline, 6 months, and 12 months.

## Results

### Gender Affirming Hormone Therapy induces progressive changes in blood DNA methylation

To investigate whether DNA methylation levels in blood change in response to GAHT, we analysed epigenome-wide methylation data in transgender women commencing feminizing hormone therapy and transgender men receiving masculinizing hormone therapy (**Fig 1A**). In both models, we profiled longitudinal samples of 13 individuals at baseline, and 6 months and 12 months GAHT (**Table 1**).

**Table 1.** Age, body mass index and baseline, 6- and 12-month biochemistry in MtF and FtM groups

|  | Transgender Women (n=13) |  |  | Transgender Men (n=13) |  |  |
| --- | --- | --- | --- | --- | --- | --- |
| Age | 29.0 (22.0, 61.0) |  |  | 23.0 (21.0, 24.0) |  |  |
| BMI | 24.2 (22.3, 24.6) |  |  | 22.8 (21.8, 30.5) |  |  |
| **Biochemistry** |  |  |  |  |  |  |
|  | Baseline | 6 months | 12 months | Baseline | 6 months | 12 months |
| FSH (IU/L) | 5.2 (2.6, 7.4) | 0.8 (0.1, 1.2) | 0.2 (0.1, 0.8) | 6.4 (3.3, 7.2) | 5.4 (4.9, 6.3) | 4.1 (3.5, 5.4) |
| LH (IU/L) | 6.1 (5.4, 6.7) | 2.0 (0.1, 3.9) | 0.1 (0.1, 2.2) | 7.5 (5.9, 8.3) | 7.2 (6.2, 10.0) | 4.7 (2.1, 7.0) |
| E2 (pmol/L) | 56.0 (50.0, 76.0) | 259.0 (196.0, 393.0) | 340.0 (294.0, 419.0) | 210.0 (90.5, 460.5) | 165.0 (92.0, 171.0) | 157.0 (120.0, 197.0) |
| T (nmol/L) | 19.6 (19.8, 20.4) | 0.7 (0.4, 1.6) | 0.5 (0.4, 0.7) | 1.6 (1.2, 1.7) | 20.9 (15.3, 27.1) | 20.4 (17.5, 29.2) |
| SHBG (nmol/L) | 51.0 (43.5, 54.0) | 74.0 (49.0, 105.0) | 76.0 (53.0, 98.0) | 51.0 (39.5, 74.5) | 37.0 (21.0, 56.0) | 33.0 (23.0, 62.0) |

Expressed as Median (interquartile range).

**Figure 1.**
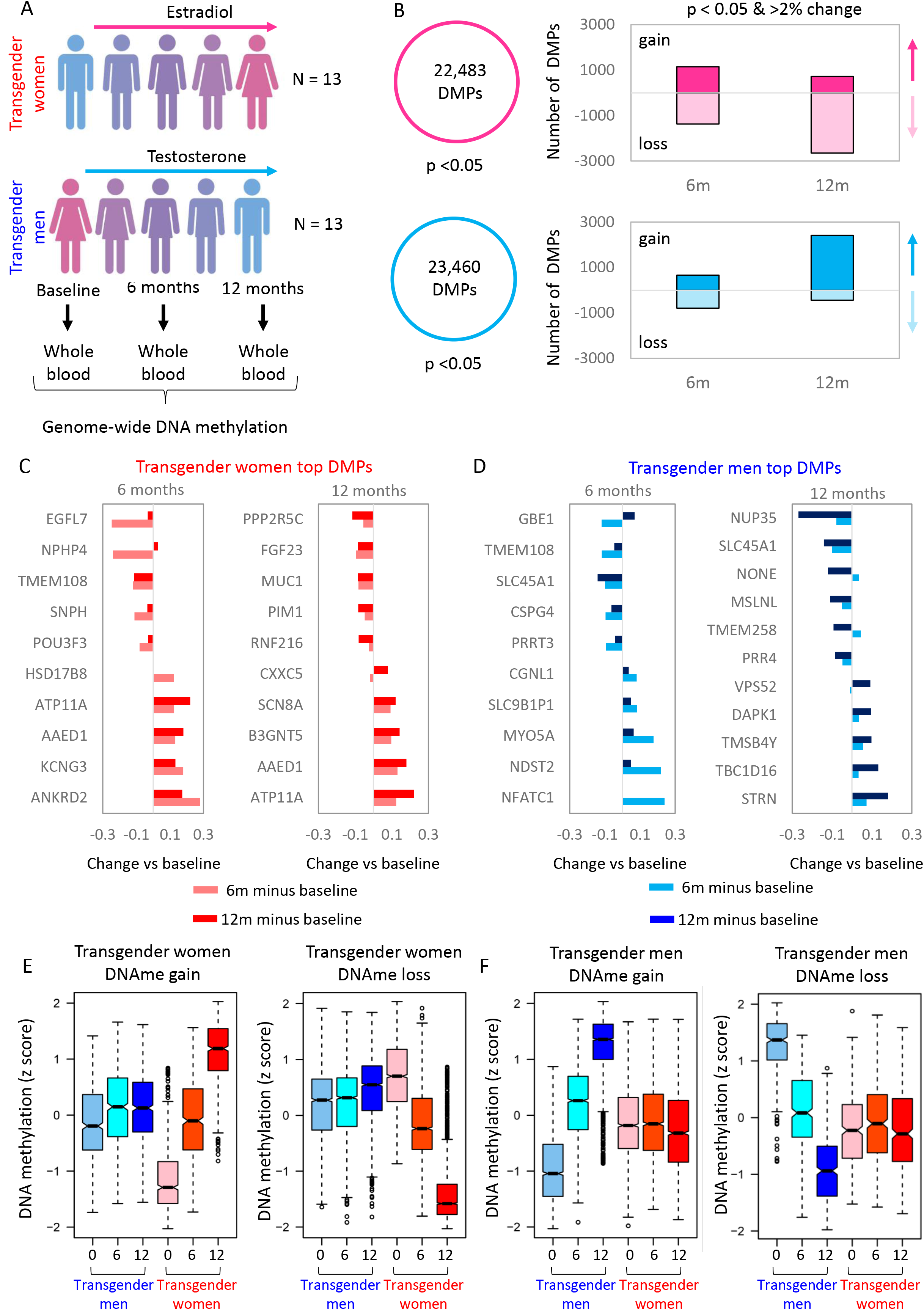
Gender Affirming Hormone Therapy model induced progressive change in blood DNA methylation. **A**. Longitudinal GAHT model with blood collection at baseline (before hormone therapy), and 6 and 12 months after treatment. In total, samples from 13 transgender women and 13 transgender men were included in the study. **B**. Number of differentially methylated probes (DMPs) with an an undajusted p-value cut-off < 0.05 and a Δβ cut-off of > 0.02. **C**. Barplot showing change in DNA methylation (Δβ) relative to baseline for top DMPs in the baseline vs 6 months or baseline vs 12 months comparison for transgender women. **D**. Barplot showing change in DNA methylation (Δβ) or top DMPs in transgender men. **E**. Boxplots of DNA methylation z-scores at DMPs that gain or lose DNA methylation at 12 months compared to baseline in transgender women. At 6 months these probes show an intermediate level of DNA methylation. **F**. Boxplots of DNA methylation z-scores at DMPs that gain or lose DNA methylation in transgender men.

In transgender women, we identified 22,483 differentially methylated probes (DMPs) after 6 months or 12 months of feminizing GAHT, with a p-value (unadjusted) < 0.05 (**Fig 1B, Table EV1**). When restricting DMPs to those that also show a greater than 2% change in methylation (mean Δβ⍰≥⍰0.02 or ≤ -0.02), we identified 2,523 DMPs after 6 months (1,151 showing gain of methylation and 1,372 showing loss of methylation), and 3,370 DMPs after 12 months (719 showing gain of methylation and 2,651 showing loss of methylation) relative to baseline (no GAHT; **Fig 1B**). In transgender men, we identified 23,460 DMPs with a p-value (unadjusted) of < 0.05 after 6 months or 12 months of masculinizing GAHT. When restricting DMPs (mean Δβ⍰>⍰0.02 or < -0.02), we identified 1,449 DMPs after 6 months (662 showing gain of methylation and 787 showing loss of methylation), and 2,758 after 12 months (2,418 showing gain of methylation and 440 showing loss of methylation) (**Fig 1B, Table EV2**). The mean methylation difference between 12 months and baseline for the top DMPs in transgender women and transgender men and the associated closest genes, are shown in **Fig 1C** and **1D**, respectively.

Next, we plotted the combined mean DMP methylation level as a z-score of masculinizing and feminizing GAHT across all groups and time-points (**Fig 1E** and **1F**). In both groups, DMPs after 12 months GAHT showed an intermediate level of methylation at the 6-month timepoint (red plots in **Fig 1E**, and blue plots in **Fig 1F**), indicating progressive change over time. The corresponding set of probes in the opposite transition group showed far less evidence of variation in response to GAHT. There was a strong correlation in Δβ between ‘baseline v 6 months’ and ‘baseline v 12 months’, with no anti-correlating DMPs (**Fig S2A** and **S2B**). Based on these DMPs the baseline group separates from the 6-month and 12-month groups, which cluster together in both transgender women and transgender men by principal component analysis (**Fig S2C** and **S2D**). In summary, DNA methylation profile is remodelled by 6 months post GAHT in a hormone-specific manner, and this pattern is maintained or enhanced by 12 months.

### Feminizing and Masculinizing GAHT remodel DNA methylation profiles in opposing directions

After 12 months GAHT, the majority of DMPs showed gains in DNA methylation in transgender men, whereas the majority of DMPs in transgender women showed loss of DNA methylation (**Fig 1B**). Only a small overlap of 64 DMPs (1.9%) was found between feminizing and masculinizing GAHT comparisons (**Fig 2A, Table S3**). Of these, 46 showed a negative correlation, with 39 gaining DNA methylation in transgender men and losing methylation in transgender women (**Fig 2B**). Clustering of individual samples by PCA based on these 39 DMPs shows a shift of samples from transgender men on masculinizing GAHT towards that of GAHT-naïve individuals assigned female at birth (down on PC2 in **Fig 2C**) and vice-versa for samples from transgender men on masculinizing GAHT (up on PC2 in **Fig 2C**). This opposing direction of DNA methylation change is also evident when clustering samples based on all 5,838 significant DMPs (unadjusted p-value < 0.05, Δβ > 0.02) (**Fig S3A**). Further, a strong correlation between ‘assigned female at birth v assigned male at birth’ Δβ and ‘baseline v 12m GAHT’ Δβ (**Fig S3B** and **S3C**), indicates that GAHT remodels DNA methylation towards the profile of the opposing sex. An example of this is an *IL21* promoter associated DMP (cg08417104) which is similarly methylated in transgender women and transgender men at baseline, but gains DNA methylation after 12 months masculinizing GAHT (mean Δβ 0.025) and loses DNA methylation after 12 months feminizing GAHT (mean Δβ -0.043) (**Fig 2D** and **2E**).

**Figure 2.**
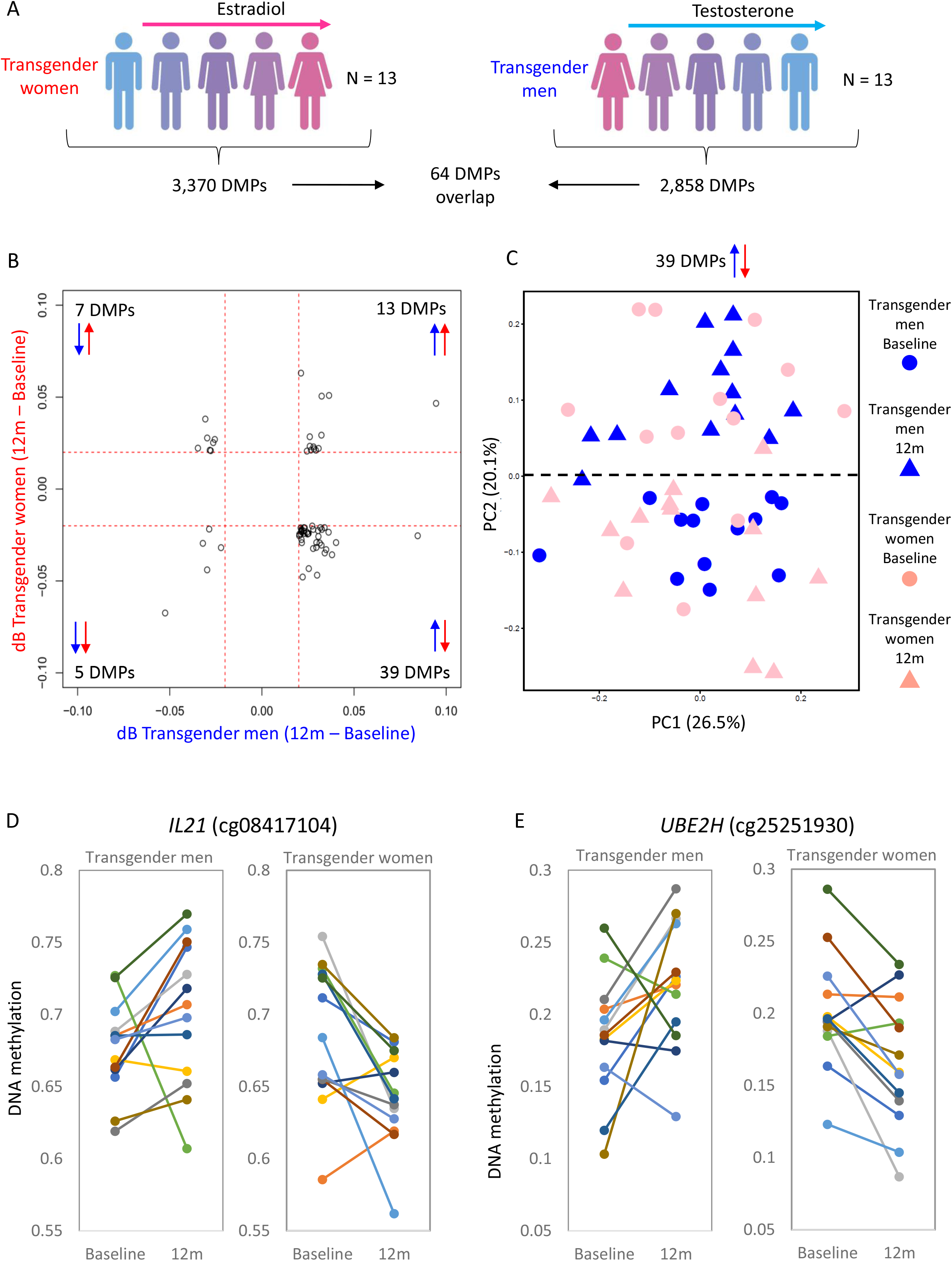
Little overlap in DMPs between transgender men on masculinizing GAHT and transgender women on feminizing GAHT. **A**. Summary of DMPs identified in transgender women and transgender men comparison at p-value < 0.05 and Δβ > 0.02. **B**. Scatter plot of change in DNA methylation during GAHT (12 months – baseline) for transgender men (x axis) and transgender women (y axis) for 64 DMPs that are significant in both GAHT groups. Green circle highlights the largest group of DMPs, 39 that show higher DNA methylation after 12 months of testosterone and lower DNA methylation after 12 months of estradiol. **C**. PCA plot of individual samples at baseline and 12 months based on the 39 anti-correlating DMPs. PC2 separates 12 months from baseline samples, with transgender women moving downwards and transgender men moving upwards. **D**. and **E**. Dot plot showing DNA methylation level for individual donors at baseline and 12 months at top probes that show an inverse change in DNA methylation between transgender women and transgender men groups. The probes are associated with *IL21* and *UBE2H* genes.

### GAHT-associates DMPs are enriched at promoters of genes associated with immunity and hormone signalling

Genes with promoter-associated DMPs, defined as a DMP within 5 kilobases of the transcription start site (TSS), were involved in numerous biological processes relating to the immune response, cell signalling, metabolism, hormone signalling, and sex-associated anatomical processes (**Fig 3 A-B, Table S4**). In transgender women receiving feminizing GAHT, 488 genes had a loss-of-methylation promoter DMPs and 196 genes had a gain-of-methylation promoter DMPs at 12 months. Enrichment of numerous immune-related biological processes including ‘immune response’, ‘response to stimulus’, ‘lymphocyte mediated immunity’, ‘positive regulation of NF-kB activity’ and ‘positive regulation of type I interferon production’, as well as ‘male genitalia development’ was observed (**Fig 3A**). Of note, genes involved in the biological process ‘male genitalia development’ included *BMP6, TBX3, LGR4, CTNNB1*, and *AR*. In transgender men on masculinizing GAHT, 104 genes were associated with loss-of-methylation promoter DMPs, and 592 genes were associated with gain-of-methylation promoter DMPs at 12 months. Enrichment of hormone signalling processes including ‘androgen receptor signalling pathway’ and ‘steroid hormone mediated signalling pathway’ was observed, including the genes *PPARGC1A, NR2E1, RWDD1*, and *DAXX* (**Fig 3B**). Additionally, ‘cell-cell signalling’, ‘response to stimulus’, ‘decidualization’, ‘interferon-gamma secretion’, and ‘response to steroid hormone’ appeared in top processes (**Fig 3B**). This indicates epigenetic remodelling at genomic loci associated with blood cell function and response to hormonal stimuli. Further, in the comparison of our DMPs to a validated pregnancy-associated 15 probe DNA methylation signature, 3/15 were present in transgender women on feminizing GAHT and none in transgender men on masculinizing GAHT, reflecting the estrogen-specific response (**Fig S4A**). Finally, we saw no overlap between our GAHT-associated DMPs and those identified in rheumatoid arthritis patients, a known sexually dimorphic autoinflammatory disease (**Fig S4B**).

**Figure 3.**
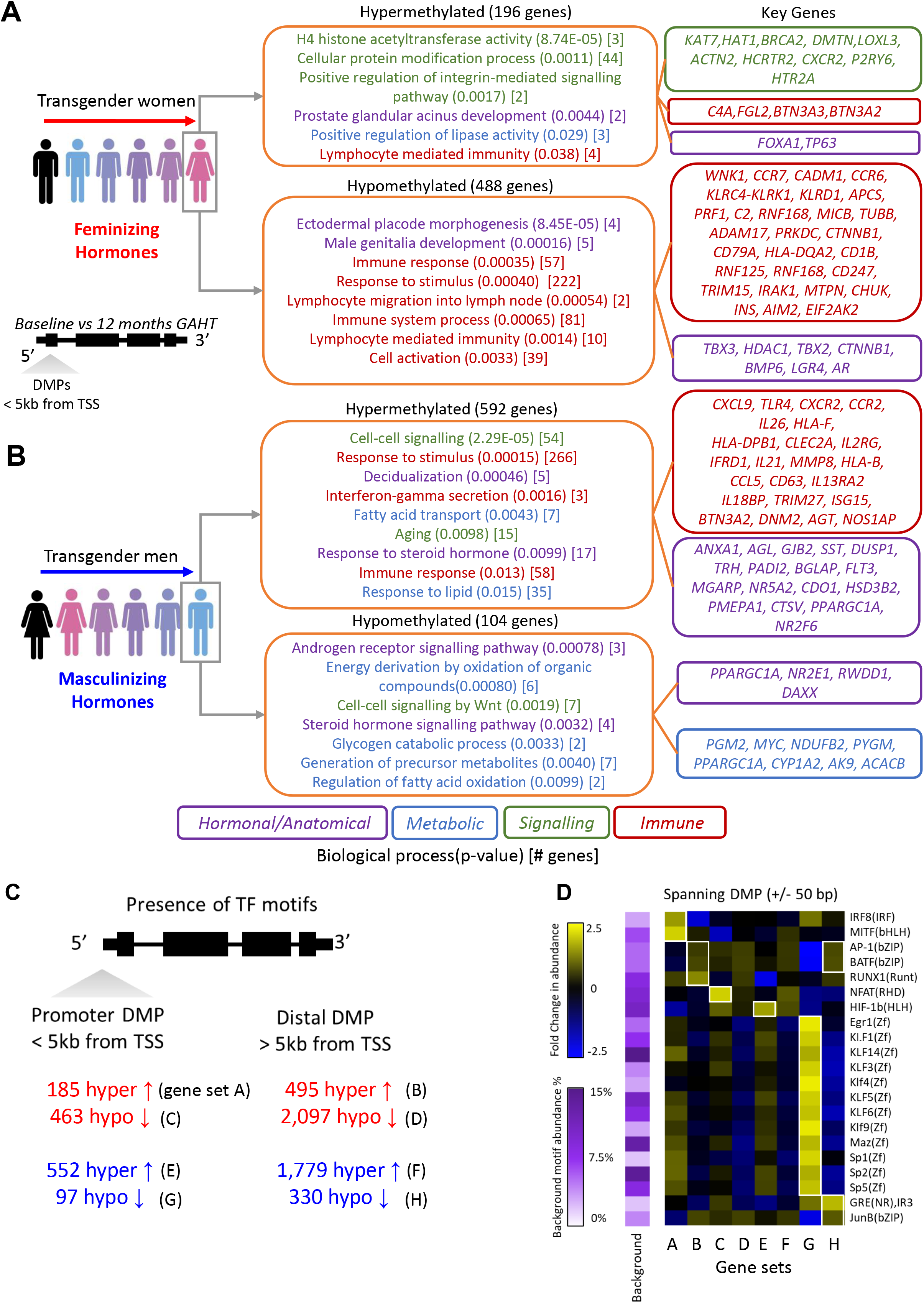
Genes with DMPs in promoters are enriched for biological processes (BP) and transcription factor binding motifs. **A**. Selected enriched BP terms of genes with a DMP (transgender women 12 months vs baseline) within 5 Kb of a transcription start site. **B**. Selected enriched BP terms of genes with a DMP (transgender men 12 months vs baseline) within 5 Kb of a transcription start site (TSS). In **A** and **B**, BP terms are followed by (p-value) and [number of target genes in term]. Key genes are highlighted on the right. **C**. Enriched TFBMs in genomic sequences spanning 50 bp upstream and downstream of promoter DMPs (PROM, < 5Kb of TSS) and distal DMPs (DIST, > 5Kb of TSS). Heatmaps show the fold change in TFBM abundance relative to average background abundance (row-scaled), with white squares indicating TFBMs with FC > 1.5 and change in abundance of > 5%.

Next, we scanned genomic regions surrounding promoter DMPs and distal DMPs for enrichment of transcription factor binding motifs (**Fig 3C**). These provide insights into the potential regulatory mechanisms at regions with dynamic DNA methylation levels following GAHT. A motif was considered enriched if it showed an increase in abundance of > 5% and a fold change in abundance of > 1.5 compared to background sequences. Generally, Egr1, Kruppel-like factor (KLF) and Sp family motifs showed higher abundance around promoter DMPs compared to distal DMPs, while AP-1 and BATF motifs were more abundant around distal DMPs (**Fig 3D**). Some motifs showed exclusive enrichment in a particular group, including IRF8 and MITF motifs in gain-of-methylation promoter DMPs in transgender women and NFAT motif in loss-of-methylation promoter DMPs in transgender women (**Fig 3D**).

### Masculinizing GAHT DMRs include an age-associated sex-specific region in the promoter of *PRR4*

Next, we examined differentially methylated regions (DMRs) which contain multiple DMPs that show correlative methylation (**Fig S5A, Table S5**). We identified three DMRs in the transgender men group at 12-months (near *PRR4, PHF19*, and *MMP17*) (**Fig S5B**), with no DMRs detected in the transgender men group at 6 months. The *PRR4* DMR was in the promoter region, overlapped an ENCODE defined open chromatin region (**Fig 4A**), and showed the largest change in DNA methylation relative to baseline, which was present at both 6 months and 12 months (**Fig S5C**). To investigate this area further, we plotted the DNA methylation levels of probes in and around the DMR (**Fig 4B**). This revealed that the region was sex-specific, showing lower DNA methylation in people assigned male at birth compared to people assigned female at birth (mean Δβ -0.12 across the DMR). Masculinizing GAHT was associated with a loss of DNA methylation (mean Δβ -0.03 across the DMR), shifting the DNA methylation pattern from that observed in people assigned female at birth towards that observed in people assigned male at birth (**Fig 4B**). Within this DMR, cg23256579 exhibited the greatest loss of methylation following masculinizing GAHT, with 12 out of 13 donors showing robust loss of methylation (mean Δβ -0.084, p-value 5.96E-05, **Fig 4C**). The promoter region of *PRR4* is known to be differentially methylated in an age-associated sex-specific manner, with people assigned male at birth starting to lose DNA methylation faster than people assigned female at birth during puberty (Inoshita et al., 2015a; McCartney et al., 2019; Suderman et al., 2017; Vershinina et al., 2021; Yusipov et al., 2020a). The sex-associated age-related methylation of cg23256579 was validated using publicly available data from another cohort (GSE131433, (Novakovic et al., 2019)) which is comprised of matched blood samples taken at birth (Guthrie cards) and in adulthood (venous blood collected at 22 to 35 years old). At this probe, we observed similar methylation levels at birth between people assigned male and people assigned female, with a loss of methylation in adult individuals assigned male at birth compared to adult individuals assigned female at birth (mean Δβ -0.142, **Fig 4D**).

**Figure 4.**
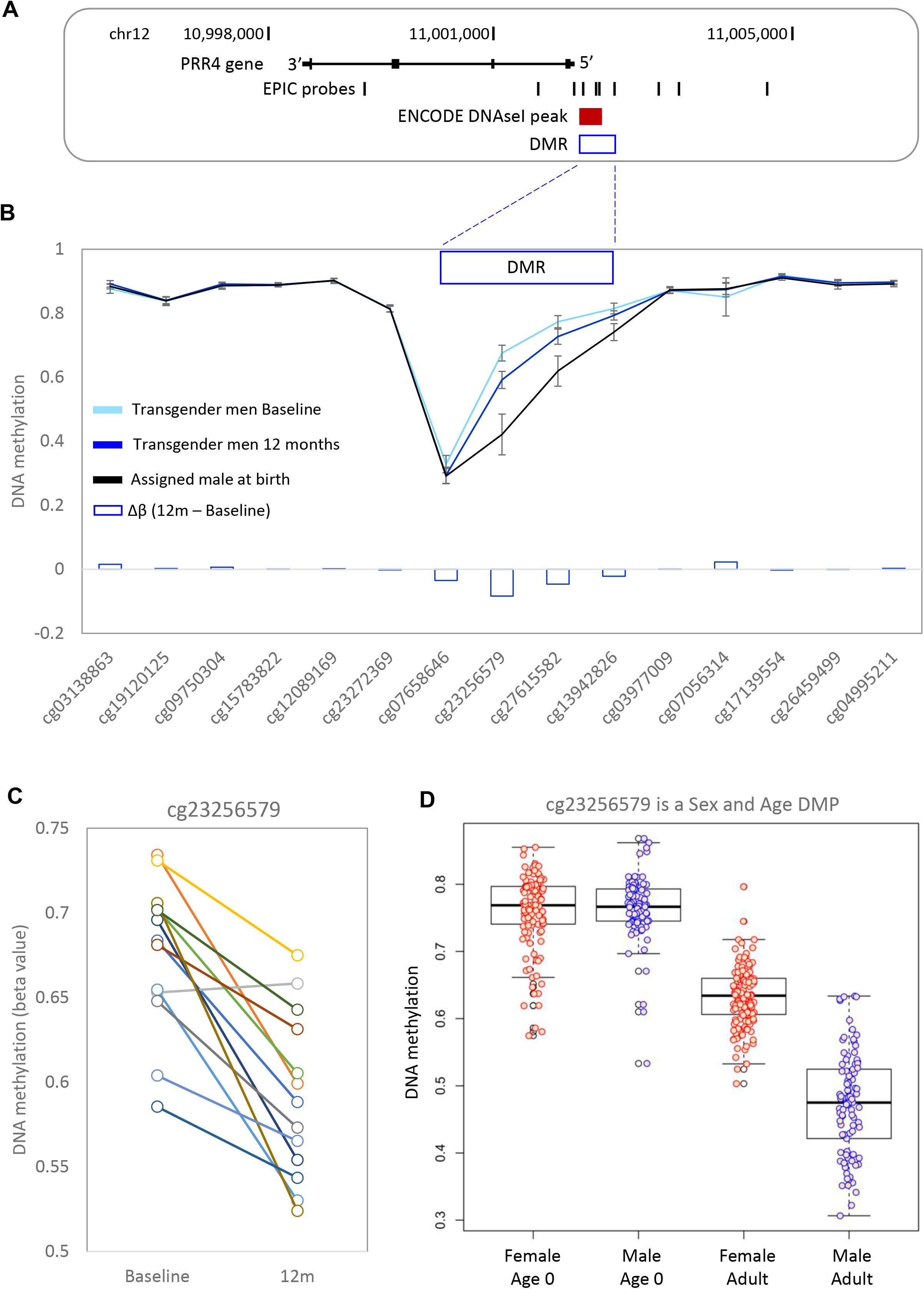
Detailed DNA methylation map of the *PPR4* gene. **A**. Map of the *PRR4* gene in hg19, showing EPIC probe locations. **B**. Mean DNA methylation level at *PRR4* for transgender men at baseline, transgender men at 12 months GAHT, and control individuals assigned male at birth. Error bars are 95% confidence intervals. Four DMPs within the DMR are losing DNA methylation in response to masculinizing GAHT, in the direction towards DNA methylation level typical of people assigned male at birth. **C**. Line plot showing individual donor changes in DNA methylation for top probe within the *PRR4* DMR - cg23256579. **D**. Boxplot of cg23256579 in blood at birth (Guthrie cards) and in adults (23-35yo), showing that sex-specific DNA methylation at this probe arises during the lifetime.

### Feminizing GAHT-associated differentially methylated regions include an age-associated sex-specific region in the promoter of *VMP1*

We identified five DMRs in transgender women at 6 months post feminizing GAHT (near *TAS1R2, ZFP36L1, MED31, VMP1*, and *SLC35D3*) and three DMRs at 12 months post feminizing GAHT (near *CAT, LANCL3*, and *HUS1*) (**Fig S5D** and **S5E**). A DMR at the 3’ UTR of *VMP1* (chr17:57,915,665-57,915,773, **Fig 5A, 5B**) showed the largest change in DNA methylation relative to baseline, with 3 probes within this DMR (cg16936953, cg12054453, and cg18942579) being significantly more methylated at both 6 months and 12 months (**Fig S3D**). By analysing the broader region at the VMP1 3’ UTR, we identified eight probes that show lower DNA methylation after 6 months of feminizing GAHT and nine showing lower DNA methylation after 12 months of feminizing GAHT compared to baseline (**Fig 5B**). The methylation levels of the top three probes in individual donors from baseline to 6 months is plotted in **Fig 5C** (mean Δβ -0.05, -0.04 and -0.03, respectively). The three probes also showed modest sex-association at baseline, with participants assigned female at birth showing lower methylation than participants assigned male at birth (mean Δβ -0.049, - 0.049, and -0.032, respectively). The sex-associated age-related methylation of two of these probes (cg16936953 and cg12054453) was validated using publicly available data (GSE131433) (**Figure 5D**), and cg12054453 was validated as a sex-associated age-related CpG showing similar methylation profiles between sexes at birth with a significant loss of methylation (p-value < 0.01, mean Δβ -0.023) in people assigned female at birth compared to people assigned male at birth by adulthood. We replicated both the *PRR4* and *VMP1* GAHT-associated DMRs in a publicly available DNA methylation dataset from an unrelated GAHT study (GSE173717; **Fig S6A-C**), strengthening our findings at these regions.

**Figure 5.**
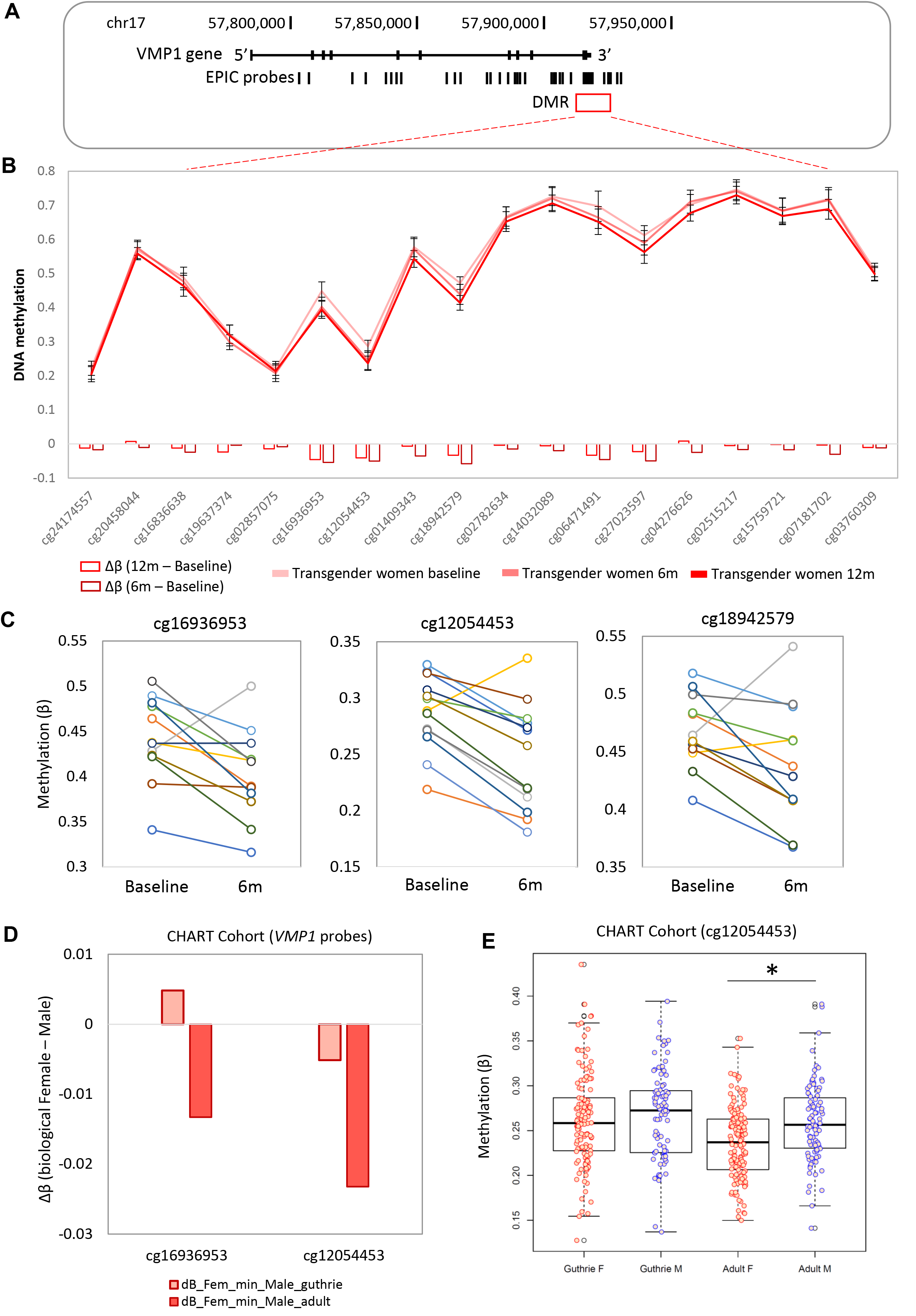
Detailed DNA methylation map of the *VMP1* gene. **A**. Map of the *VMP1* gene in hg19, showing EPIC probe locations. **B**. Mean DNA methylation level at *VMP1* for transgender women at baseline, 6 months GAHT, and 12 months GAHT. Error bars are 95% confidence intervals. Nine DMPs show the same direction of DNA methylation change within the DMR. **C**. Line plot showing individual donor changes in DNA methylation for the top 3 probes within the *VMP1* DMR. **D**. Bar plot showing difference in DNA methylation between people assigned female at birth and people assigned male at birth at the top 2 *VMP1* probes at birth and in adults. **E**. Boxplot of cg12054453 in blood at birth (Guthrie cards) and in adults (23-35yo), showing that sex-specific DNA methylation at this probe become significant during the lifetime. * p<0.01.

### X chromosome and autosomal sex-specific DNA methylation is largely unaffected by GAHT

Considering that the two top GAHT-induced DMRs, at *PRR4* and *VMP1*, were both age-associated sex-specific loci, we were interested to explore the overall effect of GAHT on sex-associated DNA methylation. By comparing GAHT-naïve transgender men and women (baseline samples), we identified 1,271 sex-associated autosomal DMPs, with 788 showing higher methylation (mean Δβ > 0.05, **Fig 6A**) and 483 showing lower methylation (Δβ < - 0.05, **Fig S7A**) in individuals assigned female at birth compared to individuals assigned male at birth (**Fig 6** and **Fig S7**). We then investigated whether these DMPs were affected by 12 months of masculinizing or feminizing GAHT, and whether this change reflected a transition towards the profile of the opposite sex. A total of 12,516 DMPs were identified on the X chromosome, of which 98.9% were unaltered after 12 months of GAHT (**Fig S7B**), indicating that GAHT has little influence on X inactivation associated DNA methylation patterns, including at the *XIST* locus (**Fig S8**). Of the 1,271 sex-associated autosomal DMPs, 1,228 (96.6%) were unaltered after 12 months of GAHT, while 19 (1.5%) were altered after feminizing GAHT and 24 (1.9%) were altered after masculinizing GAHT (**Fig 6A and Fig S7A**). Of the 43 DMPs that were affected by GAHT, 41 showed a trend towards the methylation profile of the GAHT-naïve opposite sex (**Figure S9**). Among sex-associated autosomal DMPs affected by GAHT were probes in the promoter regions of *ACACB* (cg03619736), *HLA-DPB1* (cg00798281), and *PRR4* (cg23256579 and cg27615582).

**Figure 6.**
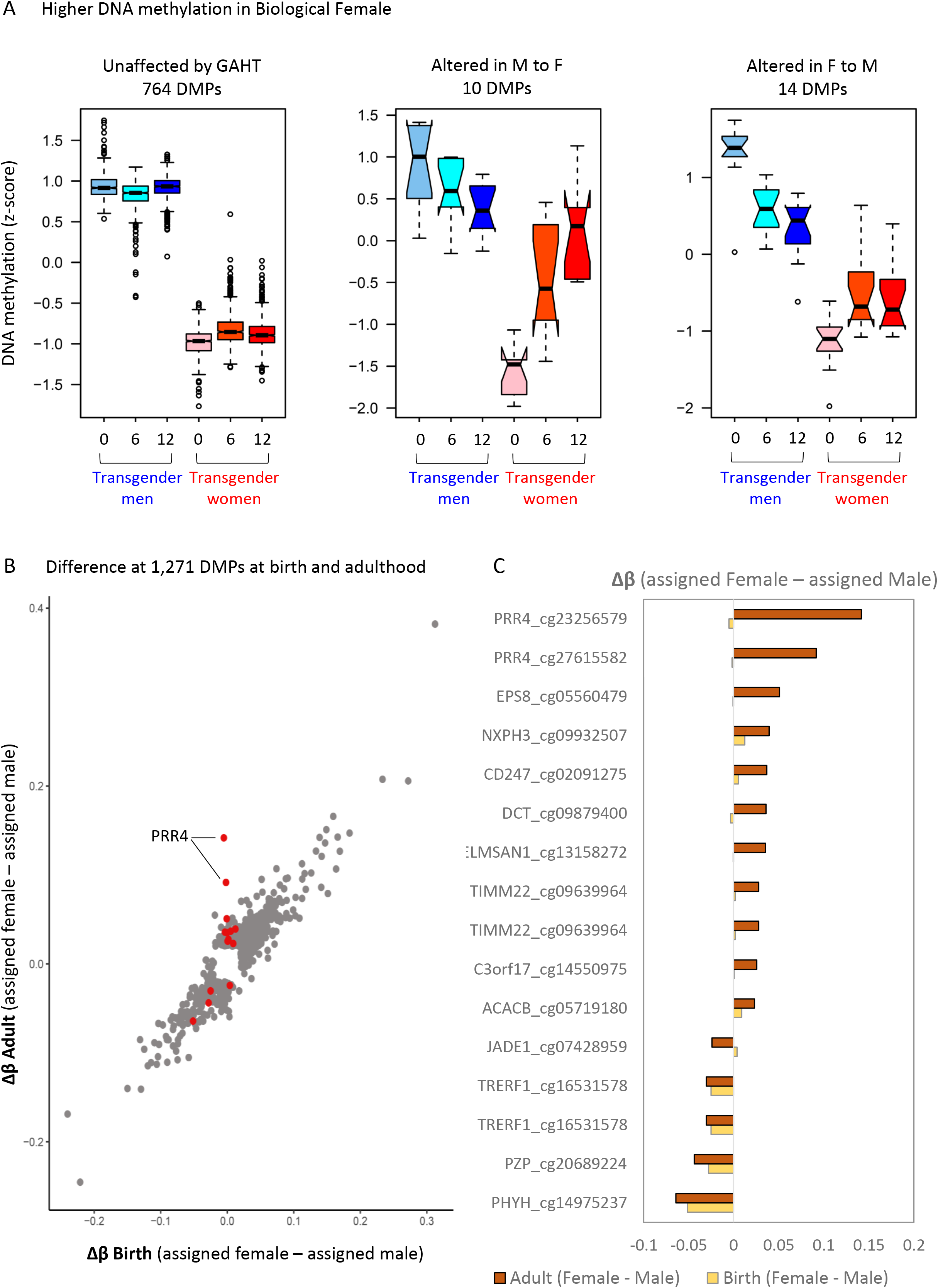
GAHT specifically influences age-associated sex-specific DMPs. **A**. Boxplot of DNA methylation (z-scored) of sex-specific autosomal DMPs that show lower DNA methylation in people assigned female at birth compared to people assigned male at birth. See Figure S5 for X chromosome and autosomal DMPs with higher DNA methylation in people assigned female at birth. 764 DMPs are not affected by GAHT, while 10 and 14 DMPs are significant in the feminizing or masculinizing GAHT comparisons. **B**. Scatter plot of change in DNA methylation (Δβ) between people assigned female at birth and people assigned male at birth at time of birth (Guthrie cards) (x axis) or in adulthood (y axis). Red dots represent probes that are significant in transgender women or transgender men comparisons, grey dots are not significant. The red dots are generally different between sexes in adults, but not at birth. **C**. Bar plot showing change in DNA methylation between sexes at probes shown in red in Fig 6B, in adults (brown) and at birth (yellow).

We then tested the hypothesis that GAHT only influenced the DNA methylation level of CpG sites that become sex-specific over time, as was the case for *PRR4* and *VMP1* loci (**Fig 4** and **Fig 5**). By correlating the change in DNA methylation between individuals assigned female at birth and individuals assigned male at birth at the 1,271 sex-associated autosomal DMPs at birth (Guthrie cards) and in adulthood, we observe a strong correlation (**Fig 6B**), indicating that most sex-specific DMPs are stable over time (grey dots). DMPs in response to GAHT are shown as red dots (**Fig 6B**), with the majority showing no evidence of sex specificity at birth, in line with them being age-associated sex-specific DMPs. This was particularly true for DMPs that lose methylation in people assigned male at birth (**Fig 6C**).

## Discussion

Gender affirming hormone therapy is a cornerstone of transgender healthcare, but it is still not known whether GAHT influences immune function, susceptibility to autoimmune disease or infection risk. With about 0.4% of the USA population identifying as transgender (Meerwijk and Sevelius, 2017), of which 80% have either taken or are considering GAHT (Nguyen et al., 2018), it is important to understand how this treatment influences the molecular biology of immune cells (Dotto, 2019). Cells of the human hematopoietic lineage are sensitive to environmental exposures, which may influence their function through epigenetic remodelling (Divangahi et al., 2021; Lau et al., 2018; Zhang and Cao, 2019). Further, there is sexual dimorphism in cytokine responses (Ter Horst et al., 2016), susceptibility to infection (Zuk, 2009), and development of autoimmune disease (Rubtsova et al., 2015). A proportion of these effects are due to hormone-associated epigenetic remodelling, with periods of hormonal change, such as puberty (Almstrup et al., 2016; Thompson et al., 2018), pregnancy (Gruzieva et al., 2019a), and menopause HRT (Bahl et al., 2015), associated with genome-wide DNA methylation changes in blood. In this study, we characterised longitudinal genome-wide DNA methylation changes in blood of transgender individuals undergoing GAHT (baseline, 6 months and 12 months post GAHT). We demonstrate that both feminizing and masculinizing GAHT alter the blood methylome in a progressive manner (**Fig 1**). This indicates that once established by 6 months, GAHT associated DNA methylation changes are stable in the presence of hormone treatment.

It remains unclear whether transgender women and transgender men on GAHT retain the autoimmunity and infection risks of their sex assigned at birth. Case reports have indicated changes in autoimmunity following commencement of GAHT, including the improvement of subacute cutaneous lupus in transgender men receiving masculinizing hormone therapy (Ocon et al., 2017) and onset of lupus or systemic sclerosis in transgender women following feminizing hormone therapy or sex reassignment surgery (Campochiaro et al., 2018; Chan and Mok, 2013; Pontes et al., 2018; Zandman-Goddard et al., 2007), suggesting a potential role of estradiol (or the absence of androgens) in the pathogenesis of autoimmune disorders. We found minimal overlap (0.1%) between GAHT-induced and the top five thousand rheumatoid arthritis-associated CpGs reported by Liu *et al*. (2013) (GEO dataset GSE42861, published in (Liu et al., 2013)) (**Supplementary Figure 7E**), and no overlap with Sjogren’s- and SLE-associated CpGs (Imgenberg-Kreuz et al., 2019) (data not shown). We did, however, find that gene promoters containing GAHT-associated DMPs are enriched for immune system processes, most markedly in genes that lose promoter DNA methylation during feminizing GAHT and genes that gain DNA methylation during masculinizing GAHT.

Previously, Aranda *et al*. (2017) reported increased methylation of the androgen receptor (*AR*) promoter in transgender women after 12 months of feminizing hormone therapy compared to baseline, and increased methylation of the estrogen receptor 1 (*ESR1*) promoter in transgender men after 6 and 12 months of masculinizing hormones therapy compared to baseline (Aranda et al., 2017). In addition, gene expression of *AR* was reduced in transgender men after masculinizing hormone therapy. We did not observe this methylation pattern in our genome-wide data. We did however identify strong DMRs at the promoter of *PRR4* and 3’ UTR region of *VMP1*, both of which we replicated in data from an unrelated GAHT cohort (**Fig 4, 5** and **EV6**).

The proline rich protein 4 (PRR4) is expressed in lacrimal glands and has been detected in a variety of other human tissues and cells (including CD4 T cells, platelets, salivary glands, and skin). The biological function of PRR4 is not known, but *PRR4* has been shown to be downregulated in cancerous laryngeal tissue and in the tear fluid of pathological conditions of the eye (Ekizoglu et al., 2018). Loss of methylation of the top probe in the DMR, cg23256579, was reported in peripheral blood cells and purified CD4+ T cells in people assigned female at birth with systemic lupus erythematosus (SLE) (Mok et al., 2016). The *PRR4* DMR exhibits an age-associated sex-specific blood methylation pattern in adults (Inoshita et al., 2015a; McCartney et al., 2019; Suderman et al., 2017; Vershinina et al., 2021; Yusipov et al., 2020a), with a recent study showing that the sex-specific loss of DNA methylation in people assigned male at birth occurs around 16 years of age (Suderman et al., 2017). This suggests that puberty-associated changes in circulating testosterone may be a potential driver of this sex-specific methylation. Our data shows that this testosterone-driven change can also be induced in adults, highlighting the epigenetic plasticity of the human immune system.

Vacuole membrane protein 1 (VMP1/TMEM49), is involved in the formation of autophagosomes and interaction between the endoplasmic reticulum and other organelles (Wang et al., 2020). Sex-specific differences in methylation at the *VMP1* DMR have been previously reported, also in an age-associated pattern (McCartney et al., 2019; Yu et al., 2021; Yusipov et al., 2020b). Suderman *et al*. (2017) found that cg12054453 and cg16936953 show significant sex differences in methylation at age 17 (people assigned female showing lower methylation) but not at birth or age seven (Suderman et al., 2017). In agreement, we found that probes within this region show modest sex association, with people assigned female at birth showing a lower methylation than people assigned male at birth. After 6 months and 12 months of feminizing GAHT, transgender women exhibit a significant loss of methylation of approximately the same magnitude as the sex difference observed at baseline, thus resembling the methylation profile of people assigned male at birth (**Fig 5**). One of the probes (cg16936953) has also been reported as progressively hypomethylated across pregnancy (Gruzieva et al., 2019a) (**Table 2**), further supporting a role of female reproductive hormones, specifically estradiol, in inducing hypomethylation in pregnancy. In recent years, this region of *VMP1* has been consistently reported as hypomethylated in blood of patients with inflammatory bowel disease (IBD), Crohn’s disease (CD) and ulcerative colitis (UC) (Adams et al., 2014; Somineni et al., 2019; Ventham et al., 2016). It is worth highlighting that the loss of methylation observed in IBD patients compared to healthy controls is more pronounced than the loss of methylation observed across feminizing GAHT (Adams et al., 2014; Ventham et al., 2016). In childhood, people assigned female at birth have been shown to have a reduced risk of CD compared to people assigned male at birth (up until age 10-14), but then an increased risk after 25 years of age (Shah et al., 2018), suggesting a potential role of reproductive hormones in the sex bias of CD. Interestingly, hypogonadism has been observed in people with IBD assigned male at birth at rates higher than expected (Swendsen, 2012), and testosterone therapy has been shown to have a positive effect in the clinical course of CD (Nasser et al., 2015). Moreover, Somineni *et al*. (2019) report that the methylation profiles of cg12054453 and cg16936953 in CD patients change after treatment to resemble methylation patterns observed in people without intestinal inflammation, and the authors conclude that CD-associated loss of methylation is likely a result of inflammation rather than CD pathogenesis. Nonetheless, the relationship between *VMP1* methylation, sex hormones, and the risk of CD warrants further investigation, as it is unclear whether transgender women are at higher risk of CD following GAHT.

Finally, the GAHT model provided a unique opportunity to determine the proportion of previously published sex-specific differences on autosomal chromosomes that are sensitive to hormonal influence post puberty. In total, GAHT affected 3% of all sex specific autosomal DMPs, but 15.8% of age-associated sex specific DMPs (**Fig 6**; Yusipov *et al*. 2020). This indicates that sex-specific DNA methylation that is present throughout life, from birth to adulthood, is hard-wired and determined by genetic and developmental programs, and not susceptible to change in response to hormones. The second conclusion from this analysis is that puberty-associated changes in DNA methylation can be induced in adults.

There are some limitations of this study that are worth highlighting. Firstly, the sample type used in this study was buffy coat, and thus potential cell-specific changes in DNA methylation are masked. We recommend future studies to investigate DNA methylation changes in particular immune cell subsets. Although the use of longitudinal samples allows for a reduced sample size, we did not have the power to look at the effect of age. Considering that we identified sex-associated age-related CpGs that are altered by GAHT, the magnitude of GAHT-induced effects may differ between age groups. It would also be informative to investigate the effect of hormone blockers in younger transgender individuals. Additionally, other molecular events, such as post-translational histone modifications and gene expression changes may be informative, especially for acute responses to GAHT. In summary, our study is the first epigenome-wide analysis of transgender men and women during GAHT, and the observed epigenetic changes in blood warrant further cell-specific functional and molecular profiling.

## Methods

### Study participants

Thirteen transgender men (assigned female at birth) seeking masculinizing GAHT and thirteen transgender women (assigned male at birth) seeking feminizing GAHT gave informed consent and consented to genetic testing of blood samples, as part of the project “The effects of cross-sex hormone therapy on bone microarchitecture in transgender individuals; a prospective controlled observational study” based at the Austin Health (Human Research Ethics Committee project HREC/17/Austin/74). Median age at the commencement of GAHT was 23 years in transgender men (IQR 21, 24)), and 29 years in transgender women (22, 61).

### Sample processing

Venous blood was collected at baseline (prior to or within 14 days of commencement of GAHT), and at 6 months and 12 months post GAHT commencement. Transgender men were taking full doses of either transdermal or intramuscular testosterone and transgender women were taking standard doses of either oral or transdermal estradiol as well as anti-androgen agents. Transgender individuals were recruited from endocrinology outpatient clinics and primary care general practice clinics specialising in transgender health in Melbourne, Australia. All 13 transgender men were receiving testosterone therapy (intramuscular (IM) testosterone undecanoate n = 6 IM testosterone enanthate n = 3, transdermal testosterone gel 1% n = 4) and all 13 transgender women were receiving estradiol (oral estradiol valerate n = 6, transdermal estradiol n = 7). A total of 77% (n = 10) of the feminizing GAHT group were taking androgen blocking therapy in addition to estradiol therapy (cyproterone acetate n = 8, cyproterone acetate and progesterone n = 1, spironolactone n = 2). No individuals in either group had undergone gonadectomy. A total of 39 longitudinal blood samples from transgender men (n = 13 at baseline, 6 months, and 12 months), and a total of 38 from transgender women (n = 13 at baseline, n = 12 at 6 months, and n = 13 at 12 months).

Part of the blood sample was sent for immediate analysis. Estradiol was measured using immunoassay (Cobas E801, Roche Diagnostics, inter-assay variation 25% at level of 100 pnmol/L or less and 25% at a level of greater than 100 pmol/L). Testosterone was measured using electrochemiluminescent immunoassay (Cobas E801, Roche Diagnostics, inter-assay CV is 5.3% at a level of 3.4 nmol/L, 4.5% at 13.3 nmol/L and 4·0% at 27·2 nmol/L). SHBG was measured on immunoassay (Cobas E801, Roche Diagnostics, inter-assay variation 6% at a level of 21 nmol/L and 6% at a level of 40 nmol/L). The remaining blood sample was centrifuged, and the plasma component removed. An aliquot of the remaining buffy coat containing concentrated circulating blood cells was stored at -30°C.

### Genomic DNA extraction and DNA methylation profiling

Buffy coats were lysed with proteinase K for 2 hours and the DNA was extracted using the Qiagen kit QIAamp® DNA Mini spin kit (Ref 56304) and eluted in 120 µL of elution buffer. Gel electrophoresis was used to confirm successful DNA extraction. Genomic DNA extracted from buffy coats was plated into 96-well plates at a concentration of 50 ng/µL (15 µL per well) and sent to Erasmus Medical Centre, Netherlands, for sodium bisulfite conversion and genome-wide methylation analysis using the Illumina Infinium Methylation EPIC Array, which measures DNA methylation across 850,000 CpG sites (referred to as EPIC ‘probes’), spanning promoter regions, gene bodies, and ENCODE-assigned distal regulatory elements.

### DNA methylation data cleaning, normalization, and statistical analysis

Raw IDAT files for were processed using the MissMethyl and minfi packages for R (Aryee et al., 2014; Phipson et al., 2016). All samples had a good quality score (mean detection p-value of <0.01). Data were normalized for both within and between array technical variation using SWAN (Subset-quantile Within Array Normalization) (Maksimovic et al., 2012). Probes with poor average quality scores (detection p-value >⍰0.01), those that overlap a SNPs at their CG site, and cross-reactive probes were removed from further analysis (Pidsley et al., 2016). This left a total of 787,296 probes for downstream analysis. Cell composition was determined using the estimateCellCounts tool, with the ‘Blood’ reference dataset (Houseman et al., 2014). Differential methylation analysis by linear regression modelling was performed using limma (Ritchie et al., 2015). Confounders and covariates were identified using principal component analysis (shown in **Figure EV1**) and were incorporated in the analysis models as required. Differentially methylated probes (DMPs) were those with an unadjusted p-value of < 0.05 and a change in methylation (delta beta or Δβ) of ≥ 2%. DMPs were assigned to the nearest gene within 1 megabase (1⍰Mb) using the GREAT tool (McLean et al., 2010). Differentially methylated regions (DMRs) were identified using the DMRcate tool (Peters et al., 2015) and Bedtools were used to intersect DMRs with individual probes (Quinlan and Hall, 2010).

### Publicly available DNA methylation data

To put our findings into context, we used previously generated EPIC array data from age-specific sex-associated profiles GSE71245 (Mamrut et al., 2015), GSE60275 (Cotton et al., 2015), GSE67393 (Inoshita et al., 2015b), 6 months of GAHT (GSE173717), and rheumatoid arthritis GSE42861 (Liu et al., 2013).

### Motif enrichment and Gene Ontology analysis

DMPs were mapped to the closest gene using GREAT (http://great.stanford.edu/public/html/) and gene promoters were scanned for enriched transcription factor binding motifs (TFBMs) using the HOMER findMotifs tool (http://homer.ucsd.edu/homer/). DNA sequences spanning DMPs (+/- 50 base pairs) were also scanned for enriched motifs using the HOMER findMotifsGenome tool. Enriched motifs were defined as those with an increased abundance of > 5%, and a fold change in abundance of > 1.5. To gain insight into biological function of genes associated with DMPs, enriched biological processes (BP), KEGG pathways, and MSigDB terms (enrichment p-value < 0.05) were also identified using HOMER.

## Data Availability

The genome-wide DNA methylation dataset produced in this study is available at Gene Expression Omnibus GSE176394

https://www.ncbi.nlm.nih.gov/geo/query/acc.cgi?acc=GSE176394

## Data Availability

https://www.ncbi.nlm.nih.gov/geo/query/acc.cgi?acc=GSE176394

## Data Availability

https://www.ncbi.nlm.nih.gov/geo/query/acc.cgi?acc=GSE176394

## Data availability

The genome-wide DNA methylation dataset produced in this study is available at Gene Expression Omnibus GSE176394 (https://www.ncbi.nlm.nih.gov/geo/query/acc.cgi?acc=GSE176394).

## Acknowledgements

BN is supported by an NHMRC (Australia) Investigator Grant (APP1173314). This project was supported by an MCRI II Theme internal funding grant.

## Author contributions

**ReS** performed analysis and wrote the original draft of the manuscript. **IB, JZ** and **AC** undertook data collection in the clinical samples and performed analysis. **AC** acquired funding and supervised the clinical study. **BW, AZ, AV**, and **ReS** performed molecular lab protocols. **KP, RS, AC, JZ** and **BN** conceptualised the study. **BN** acquired funding for the molecular analyses, performed the bioinformatics analysis and wrote the original draft of the manuscript. All authors reviewed and edited the text and approved the final manuscript.

## Conflict of interest

The authors have no conflicts of interest.

### The Paper Explained

#### Problem

In the era of personalised medicine, inclusivity of all genders and sexes in research is vital for providing equitable and inclusive healthcare. Gender affirming hormone therapy (GAHT) is a cornerstone of transgender care, yet there are no studies published to date investigating epigenome-wide changes in the blood of transgender people on GAHT. In recent years, several studies have identified a sex-specific age-related methylation signature in a cis-gendered population. It remains unclear whether transgender people on GAHT retain the DNA methylation signature of their sex assigned at birth, their affirmed gender, or a unique signature. GAHT-induced changes in DNA methylation may have implications for ‘sex-specific’ aspects of immunity such as the risk of autoimmune disorders, or the response to infection and vaccines.

#### Results

We profiled genome-wide DNA methylation in blood of transgender women and transgender men before and during GAHT (6 months and 12 months into feminizing or masculinizing hormone therapy). Feminizing and masculinizing GAHT induced differential methylation of several thousand CpG probes (DMPs) across the genome, with the majority exhibiting a progressive gain or loss of methylation over time. A small subset of DMPs were commonly altered by both masculinizing and feminizing GAHT, with the majority showing opposing effects on DNA methylation.

Genes with GAHT-associated DMPs in promoter regions were enriched for a variety sex hormone signalling pathways and immune functions, suggesting that these processes may be modulated by GAHT. Only a small percentage of the sex-specific methylation signature was altered by GAHT (approximately 3 percent), but the GAHT-induced changes consistently demonstrated a shift towards the methylation signature of the GAHT-naïve opposite sex. These included regions in *PRR4* and *VMP1*, which have been previously shown to be sex-specific and age-related in cis-gendered populations. Lastly, we compare GAHT-associated changes with those previously published in pregnancy, puberty and adolescence, and rheumatoid arthritis, which is a useful resource for follow-up studies.

#### Impact

We provide novel evidence for GAHT inducing a unique blood methylation signature in transgender people, importantly with a shift towards the methylation profile of the opposite sex. We demonstrate that a small subset of regions previously shown to exhibit robust sex-associated methylation are altered by GAHT, but that most sex-specific methylation remains stable during GAHT. This study advances our understanding of the complex interplay between sex hormones, sex chromosomes, and DNA methylation in the context of immunity. We highlight the need to broaden the field of ‘sex-specific’ immunity beyond cis-males and cis-females, as transgender people on GAHT exhibit a unique molecular profile that warrants further understanding.

#### For more Information

**Austin Health Gender Clinic**: https://www.austin.org.au/gender-clinic/

## Notes

### Competing Interest Statement

The authors have declared no competing interest.

### Funding Statement

The work was funded by an NHMRC (Australia) Investigator Grant.

### Author Declarations

Austin Health Human Research Ethics Committee (HREC/17/Austin/74)

## References

Adams, A.T., Kennedy, N.A., Hansen, R., Ventham, N.T., O⍰Leary, K.R., Drummond, H.E., Noble, C.L., El-Omar, E., Russell, R.K., Wilson, D.C., et al. (2014). Two-stage genome-wide methylation profiling in childhood-onset Crohn’s Disease implicates epigenetic alterations at the VMP1/MIR21 and HLA loci. Inflamm Bowel Dis 20, 1784–1793.

Almstrup, K., Johansen, M.L., Busch, A.S., Hagen, C.P., Nielsen, J.E., Petersen, J.H., and Juul, A. (2016). Erratum: Pubertal development in healthy children is mirrored by DNA methylation patterns in peripheral blood. Scientific reports 6, 30664.

Aranda, G., Fernández-Rebollo, E., Pradas-Juni, M., Hanzu, F.A., Kalko, S.G., Halperin, I., and Mora, M. (2017). Effects of sex steroids on the pattern of methylation and expression of the promoter region of estrogen and androgen receptors in people with gender dysphoria under cross-sex hormone treatment. The Journal of steroid biochemistry and molecular biology 172, 20–28.

Aryee, M.J., Jaffe, A.E., Corrada-Bravo, H., Ladd-Acosta, C., Feinberg, A.P., Hansen, K.D., and Irizarry, R.A. (2014). Minfi: a flexible and comprehensive Bioconductor package for the analysis of Infinium DNA methylation microarrays. Bioinformatics 30, 1363–1369.

Bahl, A., Pöllänen, E., Ismail, K., Sipilä, S., Mikkola, T., Berglund, E., Lindqvist, C., Syvänen, A., Rantanen, T., Kaprio, J., et al. (2015). Hormone Replacement Therapy Associated White Blood Cell DNA Methylation and Gene Expression are Associated With Within-Pair Differences of Body Adiposity and Bone Mass. 18, 647 –661.

Bereshchenko, O., Bruscoli, S., and Riccardi, C. (2018). Glucocorticoids, Sex Hormones, and Immunity. 9.

Bouman, A., Heineman, M.J., and Faas, M.M. (2005). Sex hormones and the immune response in humans. Human Reproduction Update 11, 411–423.

Campochiaro, C., Host, L.V., Ong, V.H., and Denton, C.P. (2018). Development of systemic sclerosis in transgender females: a case series and review of the literature. Clinical and experimental rheumatology 36 Suppl 113, 50–52.

Chan, K.L., and Mok, C.C. (2013). Development of systemic lupus erythematosus in a male-to-female transsexual: the role of sex hormones revisited. Lupus 22, 1399–1402.

Cheishvili, D., Parashar, S., Mahmood, N., Arakelian, A., Kremer, R., Goltzman, D., Szyf, M., and Rabbani, S.A. (2018). Identification of an Epigenetic Signature of Osteoporosis in Blood DNA of Postmenopausal Women. Journal of bone and mineral research : the official journal of the American Society for Bone and Mineral Research 33, 1980–1989.

Cotton, A.M., Price, E.M., Jones, M.J., Balaton, B.P., Kobor, M.S., and Brown, C.J. (2015). Landscape of DNA methylation on the X chromosome reflects CpG density, functional chromatin state and X-chromosome inactivation. Hum Mol Genet 24, 1528–1539.

Divangahi, M., Aaby, P., Khader, S.A., Barreiro, L.B., Bekkering, S., Chavakis, T., van Crevel, R., Curtis, N., DiNardo, A.R., Dominguez-Andres, J., et al. (2021). Trained immunity, tolerance, priming and differentiation: distinct immunological processes. Nat Immunol 22, 2–6.

Dotto, G.-P. (2019). Gender and sex-time to bridge the gap. EMBO Mol Med 11, e10668.

Ekizoglu, S., Ulutin, T., Guliyev, J., and Buyru, N. (2018). PRR4: A novel downregulated gene in laryngeal cancer. Oncol Lett 15, 4669–4675.

Feinberg, A.P. (2018). The Key Role of Epigenetics in Human Disease Prevention and Mitigation. N Engl J Med 378, 1323–1334.

Gabory, A., Attig, L., and Junien, C. (2009). Sexual dimorphism in environmental epigenetic programming. Molecular and Cellular Endocrinology 304, 8–18.

Gruzieva, O., Merid, S.K., Chen, S., Mukherjee, N., Hedman, A.M., Almqvist, C., Andolf, E., Jiang, Y., Kere, J., Scheynius, A., et al. (2019a). DNA Methylation Trajectories During Pregnancy. Epigenet Insights 12, 2516865719867090.

Gruzieva, O., Merid, S.K., Chen, S., Mukherjee, N., Hedman, A.M., Almqvist, C., Andolf, E., Jiang, Y., Kere, J., Scheynius, A., et al. (2019b). DNA Methylation Trajectories During Pregnancy. Epigenet Insights 12, 2516865719867090-2516865719867090.

Hall, E., Volkov, P., Dayeh, T., Esguerra, J.L., Salo, S., Eliasson, L., Ronn, T., Bacos, K., and Ling, C. (2014). Sex differences in the genome-wide DNA methylation pattern and impact on gene expression, microRNA levels and insulin secretion in human pancreatic islets. Genome Biol 15, 522.

Heard, E., Clerc, P., and Avner, P. (1997). X-chromosome inactivation in mammals. Annu Rev Genet 31, 571–610.

Houseman, E.A., Molitor, J., and Marsit, C.J. (2014). Reference-free cell mixture adjustments in analysis of DNA methylation data. Bioinformatics 30, 1431–1439.

Imgenberg-Kreuz, J., Almlof, J.C., Leonard, D., Sjowall, C., Syvanen, A.C., Ronnblom, L., Sandling, J.K., and Nordmark, G. (2019). Shared and Unique Patterns of DNA Methylation in Systemic Lupus Erythematosus and Primary Sjogren’s Syndrome. Front Immunol 10, 1686.

Inoshita, M., Numata, S., Tajima, A., Kinoshita, M., Umehara, H., Yamamori, H., Hashimoto, R., Imoto, I., and Ohmori, T. (2015a). Sex differences of leukocytes DNA methylation adjusted for estimated cellular proportions. Biology of Sex Differences 6, 11.

Inoshita, M., Numata, S., Tajima, A., Kinoshita, M., Umehara, H., Yamamori, H., Hashimoto, R., Imoto, I., and Ohmori, T. (2015b). Sex differences of leukocytes DNA methylation adjusted for estimated cellular proportions. Biol Sex Differ 6, 11.

Jones, P.A. (2012). Functions of DNA methylation: islands, start sites, gene bodies and beyond. Nat Rev Genet 13, 484–492.

Klein, S.L., and Flanagan, K.L. (2016). Sex differences in immune responses. Nature Reviews Immunology 16, 626–638.

Lau, C.M., Adams, N.M., Geary, C.D., Weizman, O.E., Rapp, M., Pritykin, Y., Leslie, C.S., and Sun, J.C. (2018). Epigenetic control of innate and adaptive immune memory. Nat Immunol 19, 963–972.

Liu, Y., Aryee, M.J., Padyukov, L., Fallin, M.D., Hesselberg, E., Runarsson, A., Reinius, L., Acevedo, N., Taub, M., Ronninger, M., et al. (2013). Epigenome-wide association data implicate DNA methylation as an intermediary of genetic risk in rheumatoid arthritis. Nat Biotechnol 31, 142–147.

Maksimovic, J., Gordon, L., and Oshlack, A. (2012). SWAN: Subset-quantile within array normalization for illumina infinium HumanMethylation450 BeadChips. Genome Biol 13, R44.

Mamrut, S., Avidan, N., Staun-Ram, E., Ginzburg, E., Truffault, F., Berrih-Aknin, S., and Miller, A. (2015). Integrative analysis of methylome and transcriptome in human blood identifies extensive sex- and immune cell-specific differentially methylated regions. Epigenetics 10, 943–957.

Markle, J.G., and Fish, E.N. (2014). SeXX matters in immunity. Trends in Immunology 35, 97–104.

McCartney, D.L., Zhang, F., Hillary, R.F., Zhang, Q., Stevenson, A.J., Walker, R.M., Bermingham, M.L., Boutin, T., Morris, S.W., Campbell, A., et al. (2019). An epigenome-wide association study of sex-specific chronological ageing. Genome medicine 12, 1.

McLean, C.Y., Bristor, D., Hiller, M., Clarke, S.L., Schaar, B.T., Lowe, C.B., Wenger, A.M., and Bejerano, G. (2010). GREAT improves functional interpretation of cis-regulatory regions. Nat Biotechnol 28, 495–501.

Meerwijk, E.L., and Sevelius, J.M. (2017). Transgender Population Size in the United States: a Meta-Regression of Population-Based Probability Samples. Am J Public Health 107, e1–e8.

Mok, A., Solomon, O., Nayak, R.R., Coit, P., Quach, H.L., Nititham, J., Sawalha, A.H., Barcellos, L.F., Criswell, L.A., and Chung, S.A. (2016). Genome-wide profiling identifies associations between lupus nephritis and differential methylation of genes regulating tissue hypoxia and type 1 interferon responses. Lupus Sci Med 3, e000183–e000183.

Moyer, A.M., Matey, E.T., and Miller, V.M. (2019). Individualized medicine: Sex, hormones, genetics, and adverse drug reactions. Pharmacol Res Perspect 7, e00541–e00541.

Nasser, M., Haider, A., Saad, F., Kurtz, W., Doros, G., Fijak, M., Vignozzi, L., and Gooren, L. (2015). Testosterone therapy in men with Crohn’s disease improves the clinical course of the disease: data from long-term observational registry study. Hormone molecular biology and clinical investigation 22, 111–117.

Nguyen, H.B., Chavez, A.M., Lipner, E., Hantsoo, L., Kornfield, S.L., Davies, R.D., and Epperson, C.N. (2018). Gender-Affirming Hormone Use in Transgender Individuals: Impact on Behavioral Health and Cognition. Curr Psychiatry Rep 20, 110.

Novakovic, B., Lewis, S., Halliday, J., Kennedy, J., Burgner, D.P., Czajko, A., Kim, B., Sexton-Oates, A., Juonala, M., Hammarberg, K., et al. (2019). Assisted reproductive technologies are associated with limited epigenetic variation at birth that largely resolves by adulthood. Nature Communications 10, 3922.

Ocon, A., Peredo-Wende, R., Kremer, J.M., and Bhatt, B.D. (2017). Significant symptomatic improvement of subacute cutaneous lupus after testosterone therapy in a female-to-male transgender subject. Lupus 27, 347–348.

Peters, T.J., Buckley, M.J., Statham, A.L., Pidsley, R., Samaras, K., r, V.L., Clark, S.J., and Molloy, P.L. (2015). De novo identification of differentially methylated regions in the human genome. Epigenetics Chromatin 8, 6.

Phipson, B., Maksimovic, J., and Oshlack, A. (2016). missMethyl: an R package for analyzing data from Illumina’s HumanMethylation450 platform. Bioinformatics 32, 286–288.

Pidsley, R., Zotenko, E., Peters, T.J., Lawrence, M.G., Risbridger, G.P., Molloy, P., Van Djik, S., Muhlhausler, B., Stirzaker, C., and Clark, S.J. (2016). Critical evaluation of the Illumina MethylationEPIC BeadChip microarray for whole-genome DNA methylation profiling. Genome Biol 17, 208.

Pontes, L.T., Camilo, D.T., De Bortoli, M.R., Santos, R.S.S., and Luchi, W.M. (2018). New-onset lupus nephritis after male-to-female sex reassignment surgery. Lupus 27, 2166–2169.

Quinlan, A.R., and Hall, I.M. (2010). BEDTools: a flexible suite of utilities for comparing genomic features. Bioinformatics 26, 841–842.

Ritchie, M.E., Phipson, B., Wu, D., Hu, Y., Law, C.W., Shi, W., and Smyth, G.K. (2015). limma powers differential expression analyses for RNA-sequencing and microarray studies. Nucleic Acids Res 43, e47.

Ronkainen, P.H., Pöllänen, E., Alén, M., Pitkänen, R., Puolakka, J., Kujala, U.M., Kaprio, J., Sipilä, S., and Kovanen, V. (2010). Global gene expression profiles in skeletal muscle of monozygotic female twins discordant for hormone replacement therapy. Aging cell 9, 1098–1110.

Rubtsova, K., Marrack, P., and Rubtsov, A.V. (2015). Sexual dimorphism in autoimmunity. J Clin Invest 125, 2187–2193.

Selmi, C., and Gershwin, M.E. (2019). Sex and autoimmunity: proposed mechanisms of disease onset and severity. Expert review of clinical immunology 15, 607–615.

Shah, S.C., Khalili, H., Gower-Rousseau, C., Olen, O., Benchimol, E.I., Lynge, E., Nielsen, K.R., Brassard, P., Vutcovici, M., Bitton, A., et al. (2018). Sex-Based Differences in Incidence of Inflammatory Bowel Diseases—Pooled Analysis of Population-Based Studies From Western Countries. Gastroenterology 155, 1079-1089.e1073.

Shepherd, R., Cheung, A.S., Pang, K., Saffery, R., and Novakovic, B. (2021). Sexual Dimorphism in Innate Immunity: The Role of Sex Hormones and Epigenetics. 11.

Short, S.E., Yang, Y.C., and Jenkins, T.M. (2013). Sex, gender, genetics, and health. Am J Public Health 103 Suppl 1, S93–S101.

Smith, Z.D., Chan, M.M., Humm, K.C., Karnik, R., Mekhoubad, S., Regev, A., Eggan, K., and Meissner, A. (2014). DNA methylation dynamics of the human preimplantation embryo. Nature 511, 611–615.

Somineni, H.K., Venkateswaran, S., Kilaru, V., Marigorta, U.M., Mo, A., Okou, D.T., Kellermayer, R., Mondal, K., Cobb, D., Walters, T.D., et al. (2019). Blood-Derived DNA Methylation Signatures of Crohn’s Disease and Severity of Intestinal Inflammation. Gastroenterology 156, 2254-2265.e2253.

Suderman, M., Simpkin, A., Sharp, G., Gaunt, T., Lyttleton, O., McArdle, W., Ring, S., Davey Smith, G., and Relton, C. (2017). Sex-associated autosomal DNA methylation differences are wide-spread and stable throughout childhood. bioRxiv, 118265.

Swendsen, C. (2012). Hypogonadism in Male Patients with Inflammatory Bowel Disease: 1540. Official journal of the American College of Gastroenterology | ACG 107.

Ter Horst, R., Jaeger, M., Smeekens, S.P., Oosting, M., Swertz, M.A., Li, Y., Kumar, V., Diavatopoulos, D.A., Jansen, A.F.M., Lemmers, H., et al. (2016). Host and Environmental Factors Influencing Individual Human Cytokine Responses. Cell 167, 1111–1124 e1113.

Thompson, E.E., Nicodemus-Johnson, J., Kim, K.W., Gern, J.E., Jackson, D.J., Lemanske, R.F., and Ober, C. (2018). Global DNA methylation changes spanning puberty are near predicted estrogen-responsive genes and enriched for genes involved in endocrine and immune processes. Clinical epigenetics 10, 62.

Ventham, N.T., Kennedy, N.A., Adams, A.T., Kalla, R., Heath, S., O’Leary, K.R., Drummond, H., Lauc, G., Campbell, H., McGovern, D.P.B., et al. (2016). Integrative epigenome-wide analysis demonstrates that DNA methylation may mediate genetic risk in inflammatory bowel disease. Nature Communications 7, 13507.

Vershinina, O., Bacalini, M.G., Zaikin, A., Franceschi, C., and Ivanchenko, M. (2021). Disentangling age-dependent DNA methylation: deterministic, stochastic, and nonlinear. Scientific reports 11, 9201.

Wang, P., Kou, D., and Le, W. (2020). Roles of VMP1 in Autophagy and ER–Membrane Contact: Potential Implications in Neurodegenerative Disorders. 13.

Weber, M., Hellmann, I., Stadler, M.B., Ramos, L., Paabo, S., Rebhan, M., and Schubeler, D. (2007). Distribution, silencing potential and evolutionary impact of promoter DNA methylation in the human genome. Nat Genet 39, 457–466.

Yu, C., Wong, E.M., Joo, J.E., Hodge, A.M., Makalic, E., Schmidt, D., Buchanan, D.D., Severi, G., Hopper, J.L., English, D.R., et al. (2021). Epigenetic Drift Association with Cancer Risk and Survival, and Modification by Sex. Cancers 13.

Yusipov, I., Bacalini, M.G., Kalyakulina, A., Krivonosov, M., Pirazzini, C., Gensous, N., Ravaioli, F., Milazzo, M., Giuliani, C., Vedunova, M., et al. (2020a). Age-related DNA methylation changes are sex-specific: a comprehensive assessment. bioRxiv, 2020.2001.2015.905224.

Yusipov, I., Bacalini, M.G., Kalyakulina, A., Krivonosov, M., Pirazzini, C., Gensous, N., Ravaioli, F., Milazzo, M., Giuliani, C., Vedunova, M., et al. (2020b). Age-related DNA methylation changes are sex-specific: a comprehensive assessment. Aging (Albany NY) 12, 24057–24080.

Zandman-Goddard, G., Solomon, M., Barzilai, A., and Shoenfeld, Y. (2007). Lupus erythematosus tumidus induced by sex reassignment surgery. The Journal of rheumatology 34, 1938–1940.

Zhang, Q., and Cao, X. (2019). Epigenetic regulation of the innate immune response to infection. Nat Rev Immunol 19, 417–432.

Zuk, M. (2009). The sicker sex. PLoS Pathog 5, e1000267.

